# Deep Learning Auto-Segmentation of Cervical Neck Skeletal Muscle for Sarcopenia Analysis Using Pre-Therapy CT in Patients with Head and Neck Cancer

**DOI:** 10.1101/2021.12.19.21268063

**Authors:** Mohamed A. Naser, Kareem A. Wahid, Aaron J. Grossberg, Brennan Olson, Rishab Jain, Dina El-Habashy, Cem Dede, Vivian Salama, Moamen Abobakr, Abdallah S.R. Mohamed, Renjie He, Joel Jaskari, Jaakko Sahlsten, Kimmo Kaski, Clifton D. Fuller

## Abstract

**Background/Purpose:** Sarcopenia is a prognostic factor in patients with head and neck cancer (HNC). Sarcopenia can be determined using the skeletal muscle index (SMI) calculated from cervical neck SM segmentations. However, SM segmentation requires manual input, which is time-consuming and variable. Therefore, we developed a fully-automated approach to segment cervical vertebra SM.

**Materials/Methods:** 390 HNC patients with corresponding contrast-enhanced computed tomography (CT) scans were utilized (300-training, 90-testing). Ground-truth single-slice SM segmentations at the C3 vertebra were manually generated. A multi-stage deep learning pipeline was developed, where a 3D ResUNet auto-segmented the C3 section (33 mm window), the middle slice of the section was auto-selected, and a 2D ResUNet auto-segmented the auto-selected slice. Both the 3D and 2D approaches trained five sub-models (5-fold cross-validation) and combined sub-model predictions on the test set using majority vote ensembling. Model performance was primarily determined using the Dice similarity coefficient (DSC). Predicted SMI was calculated using the auto-segmentation cross-sectional area. Finally, using established SMI cutoffs, we performed a Kaplan-Meier analysis to determine associations with overall survival.

**Results:** Mean test set DSC of the 3D and 2D models were 0.96 and 0.95, respectively. Predicted SMI had high correlation to the ground-truth SMI in males and females (r>0.96). Predicted SMI stratified patients for overall survival in males (log-rank p = 0.01) but not females (log-rank p = 0.07), consistent with ground-truth SMI.

**Conclusion:** We developed a high-performance, multi-stage, fully-automated approach to segment cervical vertebra SM. Our study is an essential step towards fully-automated sarcopenia-related decision-making.

## INTRODUCTION

Sarcopenia – the excessive loss of skeletal muscle (SM) mass and function – is a common and debilitating phenomenon in cancer patients [1]. While sarcopenia has been extensively studied in various cancer types, it has only recently been investigated thoroughly in head and neck cancer (HNC) [2]. Weight loss is common in HNC due to nutritional deficiencies induced by tumor geometry affecting normal tissues [3] and/or side effects caused by therapeutic interventions [4]. Although the link between treatment-associated weight loss and survival in HNC is unclear [5], sarcopenia has been strongly associated with oncologic outcomes such as overall survival and late radiation-induced toxicities [2,6,7]. Therefore, sarcopenia prediction is of paramount importance in patients with HNC.

Sarcopenia can be identified using different diagnostic criteria [8]. One quantitative method investigated in various studies is using a threshold based on the skeletal muscle index (SMI), the cross-sectional area of skeletal muscle measured on axial imaging normalized to the square of the patient’s height [9]. The SMI is most commonly calculated and referenced using computed tomography (CT) imaging of abdominal musculature [10–14]. However, abdominal imaging is not available for all HNC patients. Importantly, Olson et al. [15] and van Rijn-Dekker et al. [6] have recently suggested the C3 cervical vertebra musculature cross-sectional area may also be used to quantify sarcopenia accurately.

Current approaches to generate C3 musculature segmentations needed for SMI calculation rely on either semi-automated or completely manual segmentation [6], which can be time-consuming, introduce unnecessary errors, and suffer from interobserver variability. A fully-automated approach would be an attractive alternative to the current manual/semi-automated standard. Deep learning, which has found success in medical image segmentation [16–19], may be an ideal choice for fully-automated segmentation of SM. Several recent studies have utilized deep learning methods for automated SM measurement based on abdominal CT scans with reasonable performance [20–25]. However, to date, no studies have attempted to automate the SMI calculation workflow based on head and neck imaging.

The primary objective of this study was to develop a fully-automated approach to segment skeletal muscle at the C3 vertebral level for use in SMI calculations. These calculations could be directly used to determine sarcopenia status for predicting prognostic outcomes. To achieve this goal, we developed and implemented a two-stage deep learning system that utilizes 3D and 2D ResUNets to detect the C3 vertebra and segment the corresponding C3 musculature, respectively. We show that our approach can faithfully generate segmentations compared to the ground-truth of human-generated segmentation. By fully automating the sarcopenia determination workflow, we can ensure rapid, reproducible, and accurate measurements for use in clinical decision-making.

## MATERIALS AND METHODS

### Patient data

390 patients from the “HNSCC” publicly available dataset on The Cancer Imaging Archive (TCIA) [26–28] were selected for the analysis. The clinical and demographic characteristics of these patients are shown in **Table 1**.

**Table 1:**
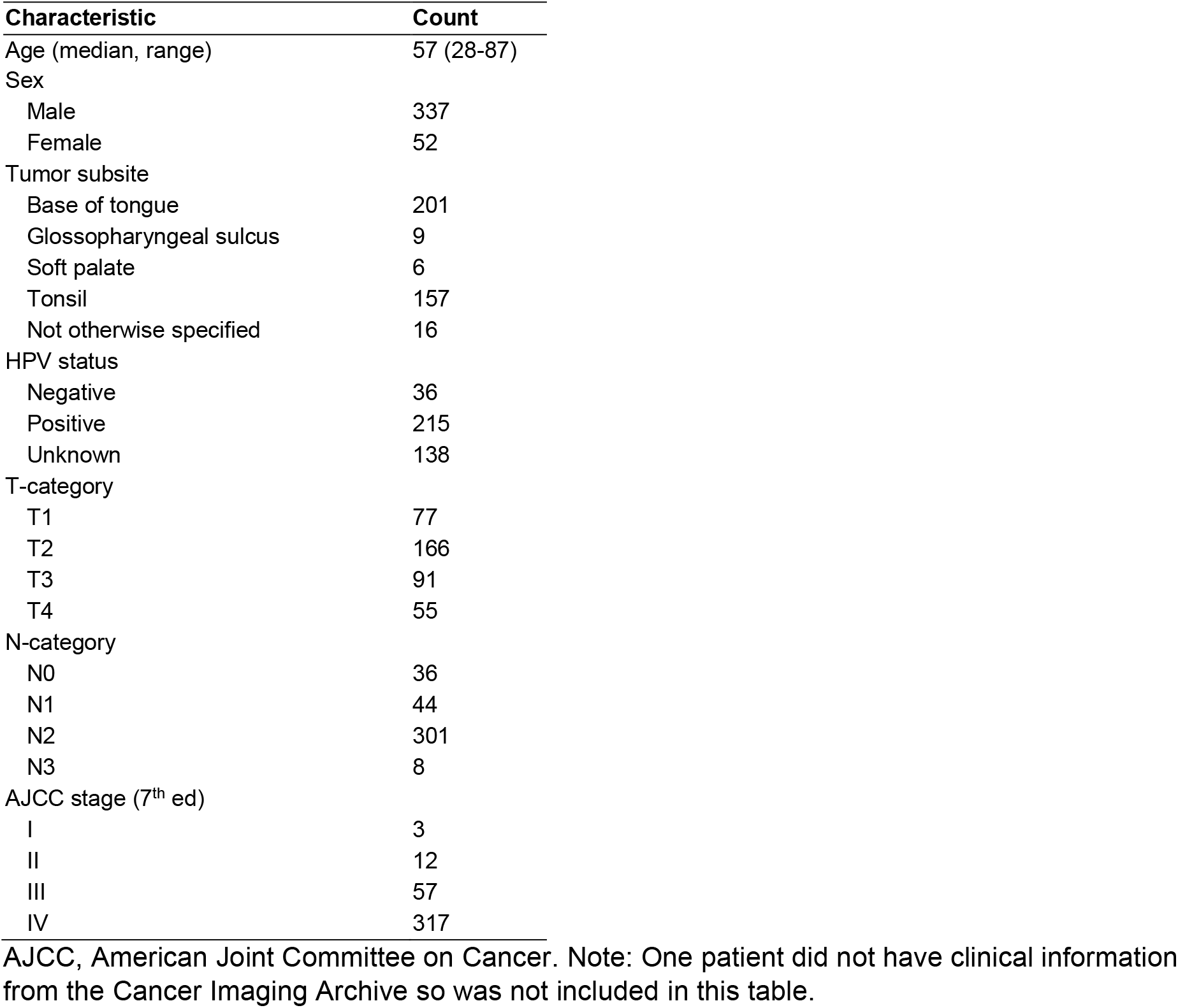
Clinical demographics of patients whose data were in this study.

### Imaging data

For the 390 patients, DICOM-formatted contrast-enhanced CT scans were acquired from the TCIA databases [26–28]. SM was manually segmented for each CT image in one slice (2D image) at the level of the C3 vertebra. The manual segmentations were performed using sliceOmatic, version 5.0 (Tomovision) using previously published Hounsfield unit thresholds to define muscle and fat [12,29]. The single-slice 2D CT images selected for the SM manual segmentation and the corresponding SM segmentations were exported as DICOM files and .tag files, respectively.

### Image Processing

The DICOM 3D volumetric and single-slice 2D CT images were converted to nifti format using the DICOM processing toolkit DICOMRTTool v. 0.3.21 [30]. The SM segmentation .tag files were converted to nifti format using an in-house Python script. The nifti files for the single-slice 2D CT images and SM segmentation were used to train the 2D segmentation model (described below). The 2D CT slice location in the C3 vertebra was extracted from the DICOM file, which was then used to generate the ground-truth segmentation mask for the C3 section, defined as a volume 33 mm in thickness centered at the location of the 2D CT slice. The tissue regions in the 3D CT images were distinguished from the background by thresholding the images using a value of greater than -500 Hounsfield units with any air gaps within the tissue region filled to generate a binary mask for the external boundaries. The generated external boundary masks and the locations of the 2D CT slices were used to create the ground-truth C3 section segmentations to train the 3D model (described below). As we have described elsewhere [31], all the images and masks were resampled to a fixed image resolution of 1 mm across all dimensions. The CT intensities were truncated in the range of [−250, 250] Hounsfield units to increase soft tissue contrast and then normalized to the range of [-1, 1] scale (**Figure 1 A,B**). We used the Medical Open Network for AI (MONAI) [32] software transformation functions to rescale and normalize images.

**Figure 1.**
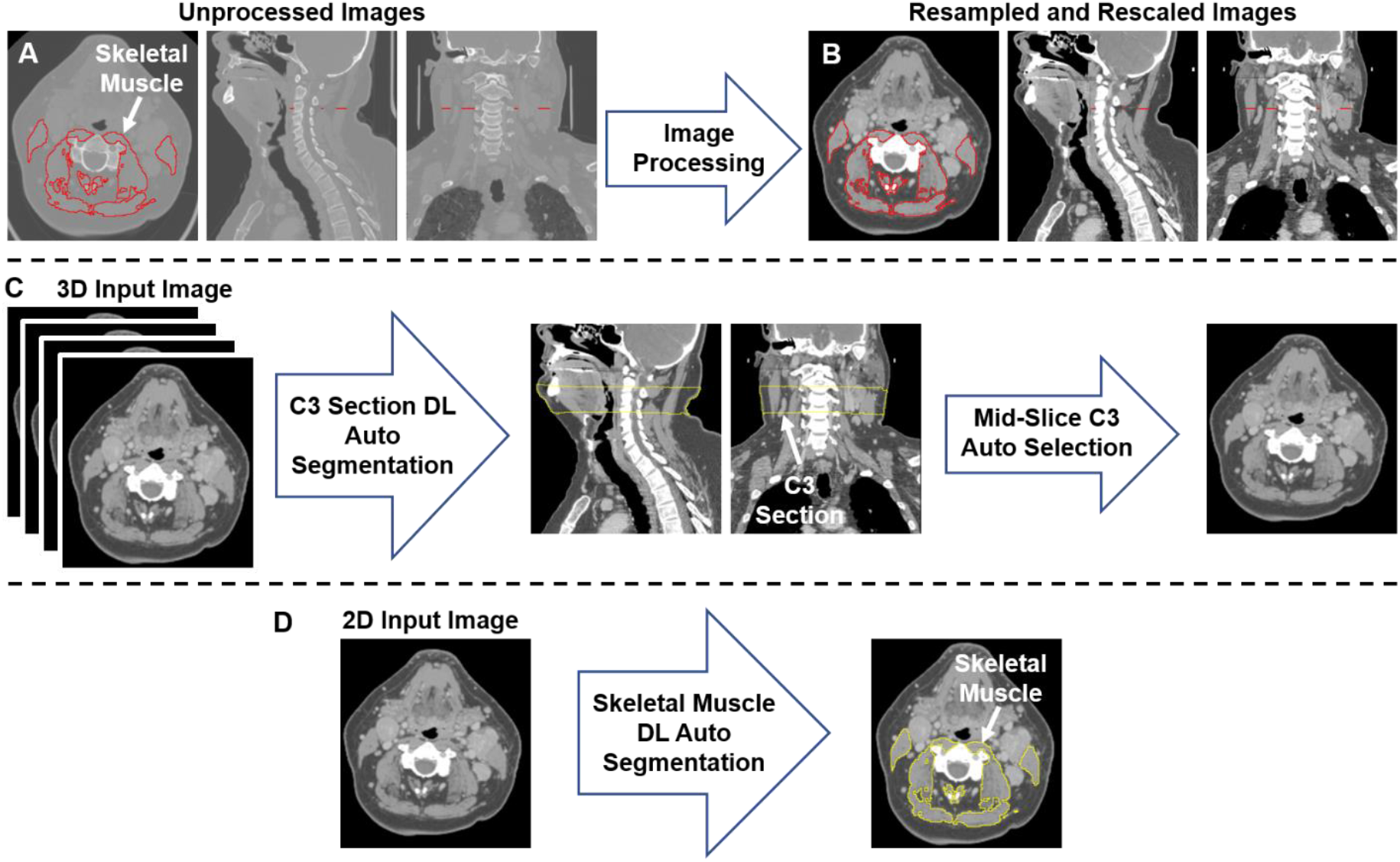
An illustration of the workflow used for skeletal muscle (SM) auto-segmentation at the C3 vertebra. (**A**) Overlays of the ground-truth SM segmentation and the original CT images. (**B**) Overlays of the ground-truth SM segmentation and the processed CT images. (**C**) An illustration of the workflow used to auto-select a single CT slice at the C3 vertebra for SM auto-segmentation. The auto-selected slice is the middle slice of the auto-segmented C3 section (33 mm in height) using a 3D ResUNet applied to the 3D volumetric CT image. (**D**) Auto-segmentation of SM using a selected C3 vertebra CT image using a 2D ResUNet model.

### Segmentation model

We used a multi-stage deep learning convolutional neural network approach for SM segmentation. Our approach was based on the UNet architecture with residual connections (ResUNet) included in the MONAI software package, as we have described in previous publications [31,33]. In the first stage of our approach (**Figure 1C**), a 3D ResUNet model auto-segmented the C3 vertebra section (33 mm), which was then followed by auto-selection of the middle slice of the section. In the second stage of our approach (**Figure 1D**), a 2D ResUNet model auto-segmented the SM on the auto-selected slice of the C3 section. Additional details of our architecture are described in **Appendix A**.

### Model implementation

We randomly split the data into 300 patients for training and 90 patients for testing. For training, we used a 5-fold cross-validation approach where the 300 patients from the training data were divided into five non-overlapping sets. Each set (60 patients) was used for model validation while the 240 patients in the remaining sets were used for training, i.e., each set was used once for testing and four times for training, leading to five sub-models. The processed CT and corresponding masks for 3D ResUNet model and 2D ResUNet models (C3 section and SM, respectively) were randomly cropped to four random fixed-sized regions (patches) of size (96, 96, 96) and (96, 96) per patch per patient, respectively. Additional details on the model implementation are described in **Appendix A**. We implemented additional data augmentation to both image and mask patches to minimize overfitting, including random horizontal flips of 50% and random affine transformations with an axial rotation range of 12 degrees and a scale range of 10%. We used the Adam optimizer for computing the parameter updates and the soft Dice loss function. The models were trained for 300 iterations with a learning rate of 2×10^−4^ for the first 250 iterations and 1×10^−4^ for the remaining 50 iterations. The values for the Adam optimizer coefficients β1 and β2 were 0.9 and 0.999, respectively. Data augmentation and loss functions were provided by the MONAI framework [32]. The final segmentations on the test set for both models were obtained by a majority vote on a pixel-by-pixel basis for all predicted segmentation masks by the 5-fold cross-validation sub-models (model ensemble), as described in a previous study [31].

### Model validation

For both the 3D ResUNet and 2D ResUNet models, we evaluated the performance on the corresponding cross-validation sets as well as the final ensemble segmentation on the test set using the Dice similarity coefficient (DSC) [34]. Specific to the 3D model, we also evaluated the accuracy of the C3 section segmentation by quantifying the absolute difference between the slice locations of the mid-section of the C3 section predicted by the 3D model and the 2D CT ground-truth image (in mm). Specific to the 2D model, we compared the SM cross-sectional areas obtained using the SM ground-truth segmentation with 1. the 2D model predicted SM segmentations on the same ground-truth CT image (Pred_GT) and 2. the 2D model predicted SM segmentations on the slices auto-selected by the 3D model (Pred_C3). We evaluated the correlation between the SM cross-sectional areas using the Pearson correlation coefficient; we also used a two-sided Wilcoxon signed-rank test to determine if these SM values were significantly different. Additionally, to derive the SMI, we normalized the SM cross-sectional areas (in cm^2^) with the patients’ heights (in m^2^). We then examined the correlation between the SMI values produced by the ground-truth and deep learning segmentations using the Pearson correlation coefficient; we also used a two-sided Wilcoxon signed-rank test to determine if these SMI values were significantly different. Based on previous work by van Rijn-Dekker et al. [6], we used **Equation 1** to calculate the cross sectional area (CSA) at the L3 lumbar level based on the CSA at the C3 cervical level and subsequently **Equation 2** to calculate the lumbar SMI:

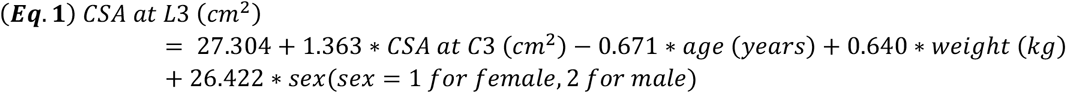

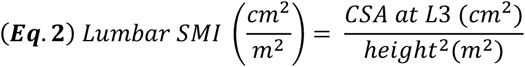

Based on previous work by Prado et al. [29], SMI thresholds of 52.4 cm^2^/m^2^ (males) and 38.5 cm^2^/m^2^ (females) were applied to lumbar SMI derived from SM ground-truth and deep learning segmentations to stratify patients by sarcopenia status (‘normal’ and ‘depleted’ muscle). These stratifications were then used for Kaplan-Meier analysis to determine associations between sarcopenia status and overall survival probabilities. To determine the sarcopenia status for the whole data set (i.e., 390 patients), we implemented Kaplan-Meier analysis on the 5-fold cross-validation data and the test data. We aggregated the SMI estimated for each cross-validation fold (i.e., 60 patients per fold) using the corresponding trained 3D and 2D models in addition to the SMI for the test data using the average predictions of the five cross-validation models.

## RESULTS

### 3D ResUNet model performance: C3 section auto-segmentation

The performance of the 3D ResUNet model for segmenting the C3 section of the neck is summarized in **Figure 2A**. When assessing the performance of each individual sub-model from our 5-fold cross-validation, the DSCs calculated between the predicted region segmentations and the ground-truth region segmentations were high and consistent between all training folds, with a mean (± standard deviation) DSC of 0.95 ± 0.01. When combining the cross-validation fold predictions using our ensemble approach, the performance on the test set increased to 0.96 ± 0.06. The middle slices of the predicted 3D regional segmentations for the test set were mostly within 4 mm of the ground-truth segmentation slice locations, with the greatest number of patients being within 1 mm (**Figure 2B**); the maximum outlier was at a distance of 10 mm. Examples of test set predictions for cases with low, medium, and high performance compared to the mean DSC are shown in **Figure 2 C-E**. As can be visually confirmed, the low-performance case still generated a segmentation such that the middle slice was contained in the C3 region.

**Figure 2.**
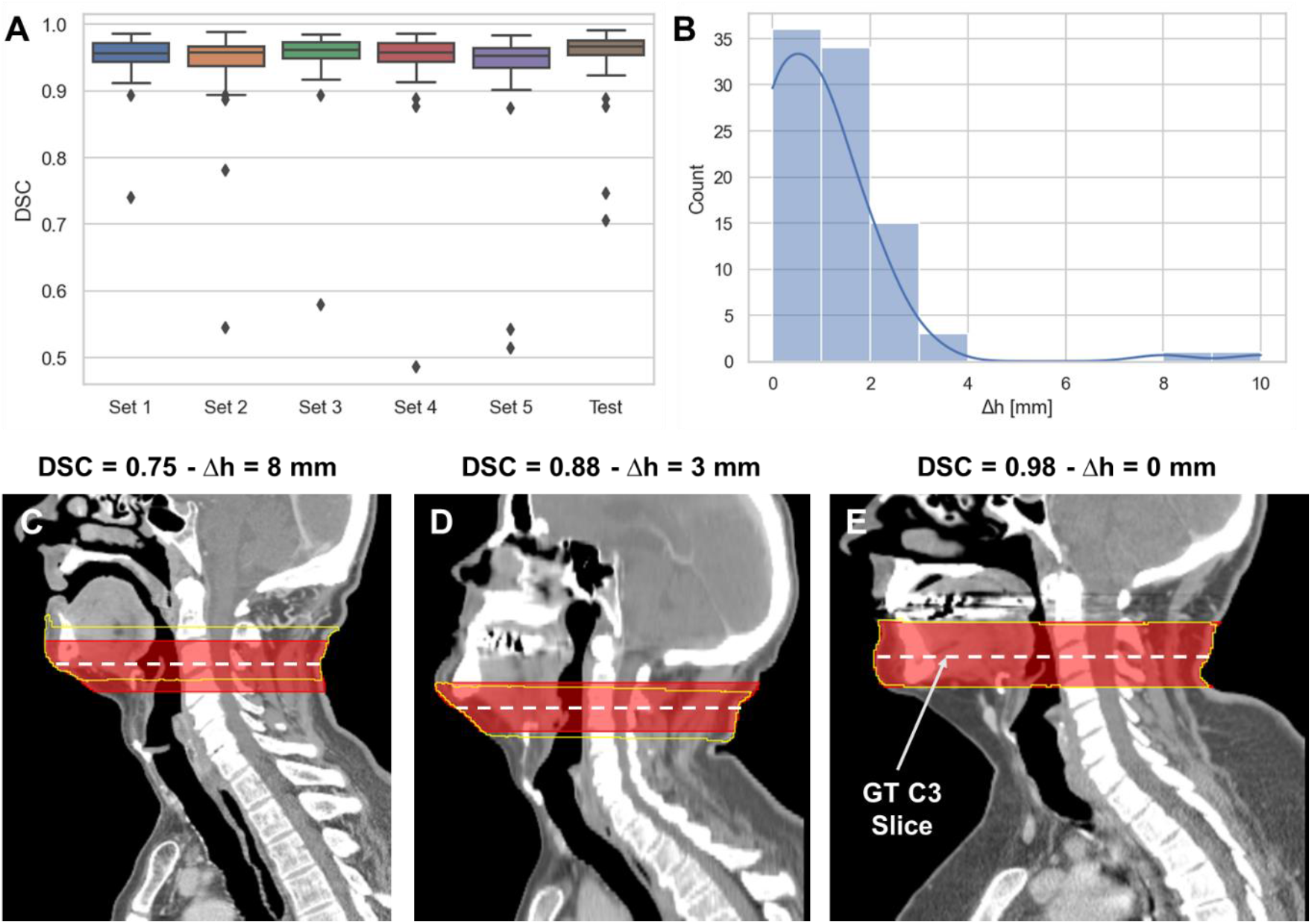
3D ResUNet model performance for segmentation of C3 vertebra section. (**A**) Boxplots of the Dice similarity coefficient (DSC) distributions for the 5-fold cross-validation data sets (Set 1 to Set 5 – 60 patients each) and the test data (90 patients). (**B**) Histogram of the absolute difference (in mm) of the C3 slice location at the middle slice of the auto-segmented C3 section and the location of the ground-truth manually segmented CT slice. Illustrative examples overlaying the C3 ground-truth segmentations (red) (33 mm centered at the ground-truth manually segmented CT slice) and predicted segmentations (yellow) on the CT images with different DSC values (low – 0.75 [**C**], medium – 0.88 [**D**], and high – 0.98 [**E**] performance compared to the mean DSC value of 0.95). The middle slice at the center of mass of the segmented C3 region was auto-selected for further skeletal muscle auto-segmentation by the 2D ResUNet model.

### 2D ResUNet model performance: SM auto-segmentation

The performance of our 2D ResUNet model for segmenting the C3 vertebra SM is summarized in **Figure 3A**. The DSCs calculated between the model-predicted segmentations and the ground-truth segmentations were high and consistent between all training folds, with a mean DSC of 0.95 ± 0.002. When combining the cross-validation fold predictions using our ensemble approach, the mean DSC performance on the test set remained consistent at 0.95 ± 0.02. The cross-sectional areas derived from the 2D model predictions using both the ground-truth slice locations and auto-selected slice locations from the 3D ResUNet model were highly correlated to the cross-sectional areas derived from the ground-truth segmentations (**Figure 3B**). The predicted areas using the ground-truth slice locations had a Pearson r=0.98 (p < 0.0001) and nonsignificant Wilcoxon test (p=0.43). Similarly, the predicted areas using the auto-selected slice locations had a Pearson r=0.98 (p < 0.0001) and nonsignificant Wilcoxon test (p=0.22). Examples of test set predictions for cases with low, medium, and high performance compared to the mean DSC for predictions using ground-truth slice location are shown in **Figure 3 C-E**. As can be visually confirmed, the low-performance case successfully generated a segmentation for musculature that was not included in the ground-truth segmentation. Moreover, the predictions using the auto-selected slice location from the 3D ResUNet model yielded virtually indistinguishable results for the low-performance and medium-performance cases (**Figure 3 F,G)** and identical results for the high-performance case (**Figure 3E**). Finally, when investigating the percentage difference in cross-sectional areas between the model-generated and ground-truth segmentations, there was no significant difference when using the ground-truth slice location or the auto-selected slice location (p=0.37) (**Figure 3H**).

**Figure 3.**
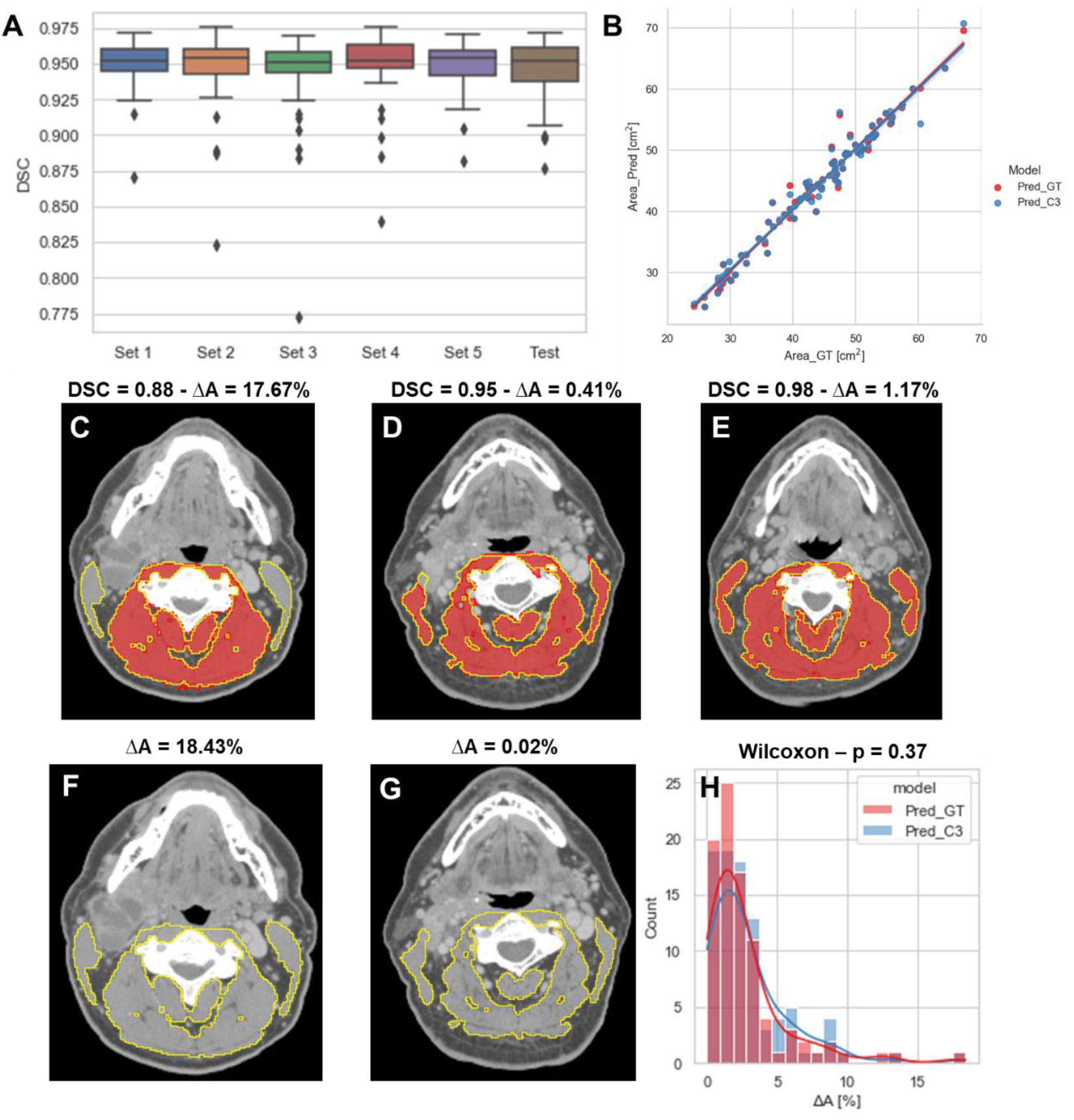
2D ResUNet model performance for segmentation of C3 skeletal muscle (SM). (**A**) Boxplots of the Dice similarity coefficient (DSC) distributions for the 5-fold cross-validation datasets (Set 1 to Set 5 – 60 patients each) and the test data (90 patients). (**B**) A scatter plot of the SM cross-sectional area using the ground-truth manual segmentation (x-axis) and the SM cross-sectional areas (y-axis) using predicted segmentations of the 2D ResUNet applied to the ground-truth CT image slice (Pred_GT) and the auto-selected CT image slice using the C3 section auto-segmentation (Pred_C3). Illustrative examples overlaying the skeletal muscle (SM) ground-truth segmentations (red) and predicted segmentations (yellow) on the same ground-truth CT images (**C, D, E**) and auto-selected CT images (**F, G**) with different DSC values (low – 0.88, medium - 0.95, and high – 0.98 compared to the mean estimated DSC value of 0.95). The auto-selected CT image for the high-performance example was identical to the ground-truth image and therefore provided the same segmentation as shown in panel C. (**H**) Histogram of percentage difference of SM cross-sectional areas between ground-truth segmentations compared to the predicted SM cross-sectional areas (ΔA%) corresponding to the model using ground-truth slice location (red) or auto-selected slice location (blue).

### SMI measurement comparisons

We compared SMI values for test set patients calculated using ground-truth SM segmentations with predicted SMI values calculated using SM segmentations generated from our 2D ResUNet models using the ground-truth slice location (**Figure 4A**) or auto-selected slice location (**Figure 4B**). Both model SM segmentations led to predicted SMI values that were highly correlated to the ground-truth SMI values. The predicted SMI values using the ground-truth slice location for males and females both had a Pearson r=0.98 (p < 0.0001) and nonsignificant Wilcoxon signed-rank tests (p=0.17 and p=0.43, respectively) compared to ground-truth SMI values. Similarly, the predicted SMI values using the auto-selected slice location for males and females had Pearson r values of 0.97 and 0.96, respectively (both p < 0.0001) and nonsignificant Wilcoxon signed-rank tests (p=0.19 and p=0.98, respectively) compared to the ground-truth SMI values.

**Figure 4.**
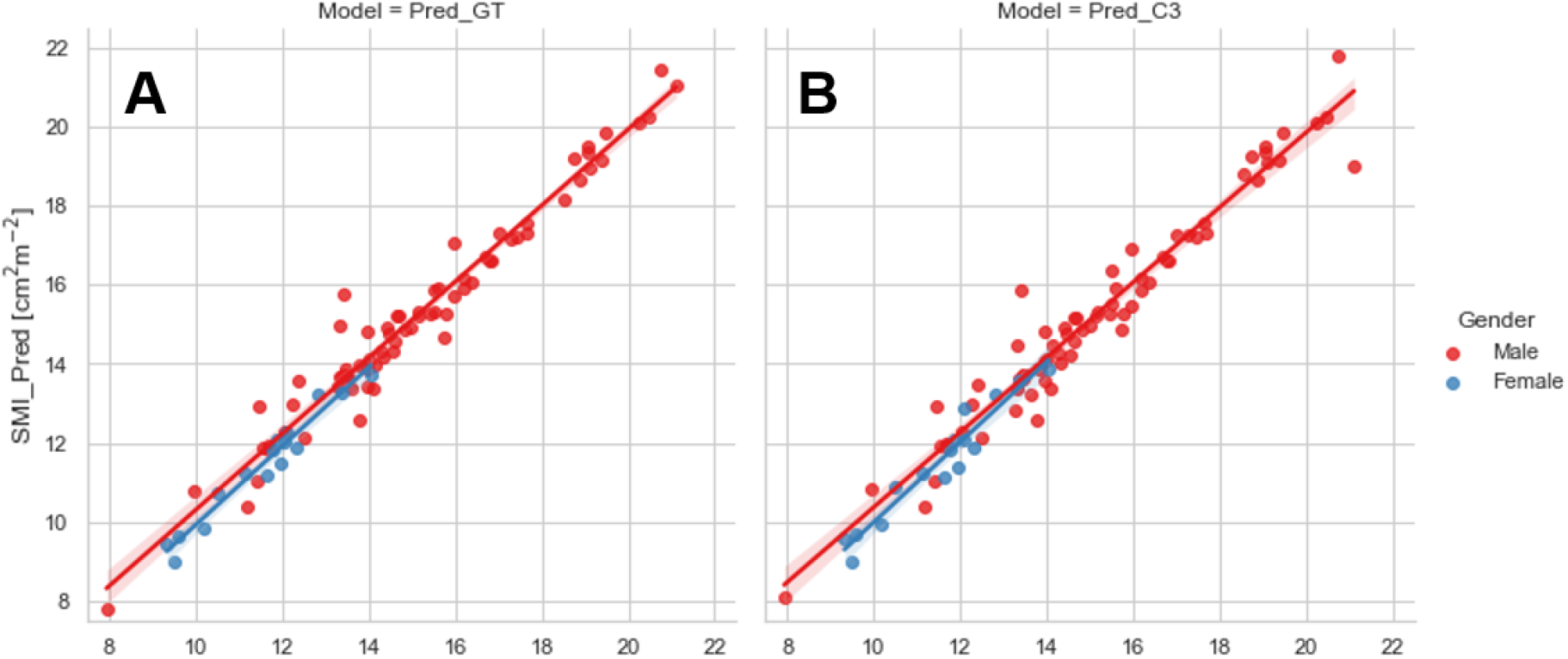
Scatter plots of the skeletal muscle index (SMI) values determined for test set patients (stratified by gender) using the ground-truth manual segmentation (x-axis) and those determined using predicted segmentations of the 2D ResUNet (y-axis) using (**A**) the ground-truth CT image slice (Pred_GT) and (**B**) the auto-selected CT image slice using the C3 section auto-segmentation (Pred_C3).

### Survival analysis

The results of the overall survival analysis based on sarcopenia thresholds are shown in **Figure 5**. Independent of the method of SMI calculation (GT, Pred_GT, or Pred_C3), there were significant differences in overall survival of males between those with normal and depleted muscle tissue (**Figure 5 A-C**), while females exhibited no significant difference (**Figure 5 D-F**). Hazard ratios (95% confidence intervals) in males for GT, Pred_GT, and Pred_C3 were 1.82 (1.1-3.0), 1.95 (1.18-3.22), and 1.97 (1.19-3.25), respectively. Hazard ratios (95% confidence intervals) in females for GT, Pred_GT, and Pred_C3 were 2.76 (0.59-13.02), 3.4 (0.73-15.83), and 3.72 (0.8-17.31), respectively.

**Figure 5.**
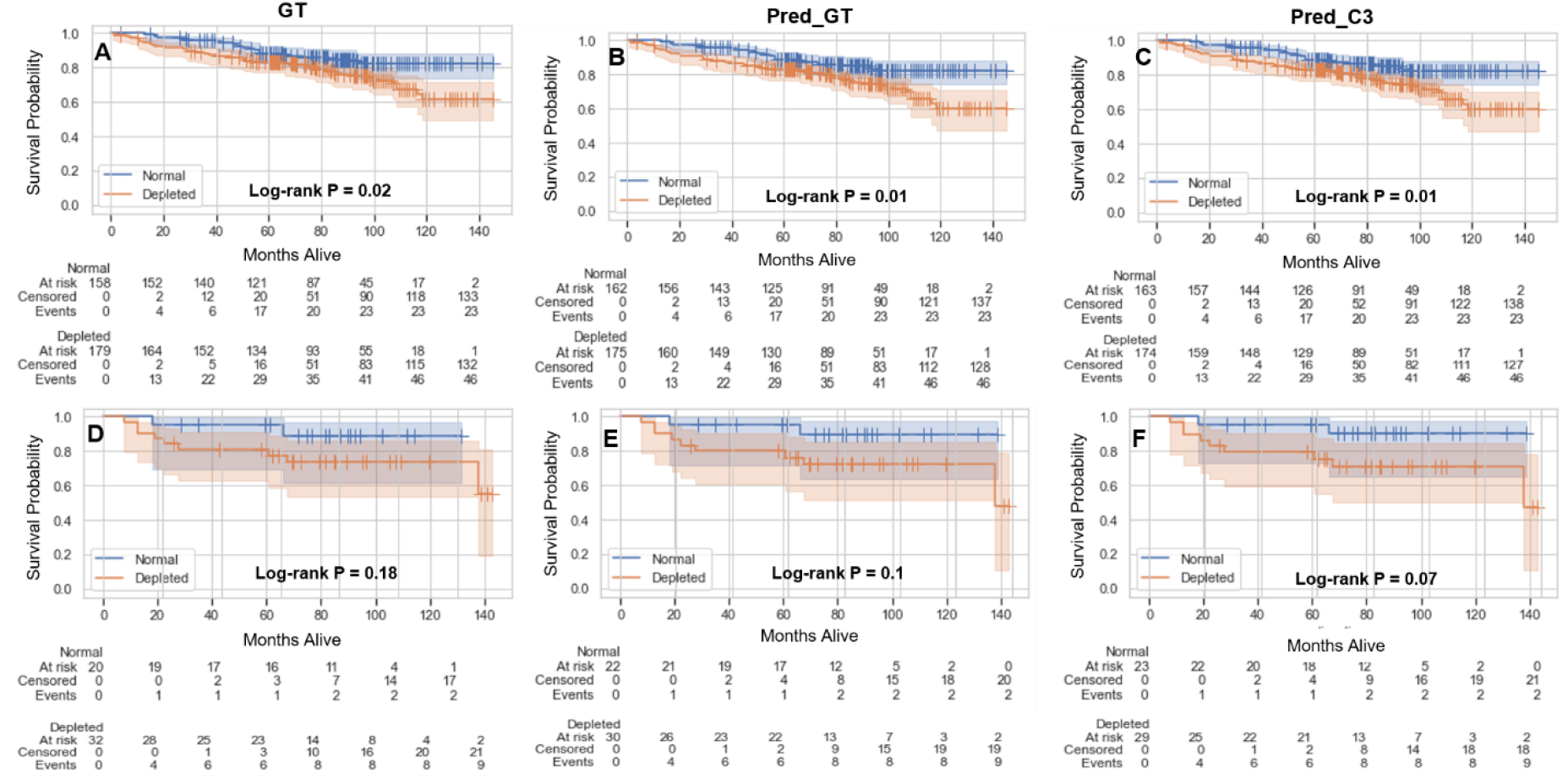
Kaplan-Meier plots showing overall survival probabilities (test and validation set combined, 390 patients) as a function of time in days for estimated skeletal muscle (SM) index (normal vs depleted) in male (**A-C**) and female (**D-F**) patients using the ground-truth SM segmentation (GT) (**A, D**), auto-segmented SM using the ground-truth slice location (Pred_GT) (**B, E**), and auto-segmented SM using the auto-selected slice location (Pred_C3) (**C, F**).

## DISCUSSION

In this study, we have utilized a multi-stage deep learning approach to segment the C3 region of the head and neck, auto-select a single representative slice, and auto-segment the corresponding SM. Our approach determined slice location and segmented SM with high accuracy when compared to ground-truth segmentations. By fully automating this workflow, we have enabled more rapid testing and application of sarcopenia-related clinical decision-making. To our knowledge, this is the first study to fully automate sarcopenia prediction based on non-abdominal imaging.

We utilized both 2D and 3D ResUNet models in our approach. By decomposing the C3 detection and SM segmentation problem into two separate tasks, we ensure that accurate representations of patient anatomy are identified by the models (C3 region) and subsequently maximize performance for SM segmentation. While previous SM auto-segmentation studies often required specific slices as model inputs [20,25] or utilized separate pre-processing software [22,24], multi-stage deep learning methods have recently been adapted in this domain as well [21,23]. Both the 2D and 3D ResUNet models that make up our segmentation pipeline had high performance, with mean DSC values in the test set above 0.95. Importantly, the performance of our C3 SM segmentation model is comparable to that of previous L3 SM deep learning segmentation models, which also demonstrate test set DSCs of ∼0.95 [20–25]. Moreover, for cases with relatively low performance, we visually confirmed that results were reasonable, i.e., the auto-selected slice was still contained within the C3 region for the 3D model, and the correct musculature was segmented on the 2D model. Importantly, we also showed minimal differences in the 2D SM segmentation model regardless of how the slice location was determined, indicating the model is robust to the specific C3 slice location. Consistent with quantitative measures of segmentation performance, using our deep learning segmentations to calculate SMI demonstrated a high correlation with ground-truth SMI independent of gender stratification.

Several previous studies have demonstrated that SMI values can be used to stratify patients into subgroups that are strongly associated with prognostic outcomes [2,6,7]. Using lumbar SMI conversion equations previously derived by van Rijn-Dekker et al. [6] and validated SMI thresholds [12], we demonstrated that calculations based on our deep learning segmentations predict similar overall survival outcomes as calculations based on ground-truth segmentations. Moreover, p-values for all methods were significant for males but not females. These results are consistent with recent literature by Olson et al. [15] which emphasized that sarcopenia was associated with poor survival outcomes in males but not in females. Our results suggest that our automated methods are dependable for use in prognostic outcome prediction.

While our study presents encouraging results towards full automation of sarcopenia-related clinical decision-making for HNC patients, there were some limitations. First, we only tested our method on pre-therapy images. Importantly, some studies have suggested prognostic evidence is higher for post-therapy sarcopenia than pre-therapy sarcopenia in HNC [7]. Therefore, further confirmatory work is needed to ensure our methods can be used accurately and reproducibly for post-therapy imaging. Moreover, we have limited our analysis to CT images as CT is the most common imaging modality for HNC radiotherapy treatment planning. However, the use of additional modalities for SM segmentation, i.e., magnetic resonance imaging, as has been utilized in other studies [35–37], may warrant additional auto-segmentation investigations. Finally, while we believe current model performance is satisfactory for clinical applications as demonstrated by comparisons with ground-truth segmentations and SMI measures, different architectural choices or ensemble approaches could be further explored to improve performance.

## CONCLUSIONS

In summary, using open-source toolkits and public data, we applied 3D and 2D deep learning approaches to head and neck CT images to develop an end-to-end automated workflow for SM segmentation at the C3 vertebral level. When evaluated on independent test data, our fully-automated approach yielded mean DSCs of up to 0.96 for segmenting the C3 vertebra region and 0.95 for segmenting the corresponding SM. Cross-sectional areas and calculated SMI values derived from our approach were highly correlated to ground-truth (r>0.95), indicating their potential clinical acceptability. Moreover, our methods can be reliably combined with validated SMI thresholds for use in prognostic stratification. Our study is an essential first step towards fully-automated workflows for sarcopenia-related clinical-decision making. Future studies should consider incorporating additional imaging timepoints and modalities for automated sarcopenia prediction.

## Data Availability

All raw imaging data are available online at: https://wiki.cancerimagingarchive.net/display/Public/HNSCC. Segmentation data are avaliable upon reasonable request to the authors.

## Abbreviations

CT: (computed tomography)
DSC: (Dice similarity coefficient)
HNC: (head and neck cancer)
ROI: (region of interest)
SM: (skeletal muscle)
SMI: (skeletal muscle index)

## ACKNOWLEDGMENTS

The authors acknowledge Sunita Patterson (Research Medical Library, MD Anderson Cancer Center) for editorial assistance.

## REFERENCES

[1] Anjanappa M, Corden M, Green A, Roberts D, Hoskin P, McWilliam A, et al. Sarcopenia in cancer: Risking more than muscle loss. Tech Innov Patient Support Radiat Oncol 2020;16:50–7.

[2] Surov A, Wienke A. Low skeletal muscle mass predicts relevant clinical outcomes in head and neck squamous cell carcinoma. A meta analysis. Ther Adv Med Oncol 2021;13:17588359211008844.

[3] Zhao J-Z, Zheng H, Li L-Y, Zhang L-Y, Zhao Y, Jiang N. Predictors for weight loss in head and neck cancer patients undergoing radiotherapy: a systematic review. Cancer Nurs 2015;38:E37–45.

[4] Powrózek T, Dziwota J, Małecka-Massalska T. Nutritional Deficiencies in Radiotherapy-Treated Head and Neck Cancer Patients. J Clin Med 2021;10:574.

[5] Ghadjar P, Hayoz S, Zimmermann F, Bodis S, Kaul D, Badakhshi H, et al. Impact of weight loss on survival after chemoradiation for locally advanced head and neck cancer: secondary results of a randomized phase III trial (SAKK 10/94). Radiat Oncol 2015;10:1–7.

[6] van Rijn-Dekker MI, van den Bosch L, van den Hoek JGM, Bijl HP, van Aken ESM, van der Hoorn A, et al. Impact of sarcopenia on survival and late toxicity in head and neck cancer patients treated with radiotherapy. Radiother Oncol 2020;147:103–10.

[7] Findlay M, White K, Stapleton N, Bauer J. Is sarcopenia a predictor of prognosis for patients undergoing radiotherapy for head and neck cancer? A meta-analysis. Clin Nutr 2021;40:1711–8.

[8] Han A, Bokshan SL, Marcaccio SE, DePasse JM, Daniels AH. Diagnostic criteria and clinical outcomes in sarcopenia research: a literature review. J Clin Med 2018;7:70.

[9] Hua X, Liu S, Liao J-F, Wen W, Long Z-Q, Lu Z-J, et al. When the loss costs too much: a systematic review and meta-analysis of sarcopenia in head and neck cancer. Front Oncol 2020;9:1561.

[10] Cho Y, Kim JW, Keum KC, Lee CG, Jeung HC, Lee IJ. Prognostic significance of sarcopenia with inflammation in patients with head and neck cancer who underwent definitive chemoradiotherapy. Front Oncol 2018;8:457.

[11] Stone L, Olson B, Mowery A, Krasnow S, Jiang A, Li R, et al. Association between sarcopenia and mortality in patients undergoing surgical excision of head and neck cancer. JAMA Otolaryngol Neck Surg 2019;145:647–54.

[12] Grossberg AJ, Chamchod S, Fuller CD, Mohamed ASR, Heukelom J, Eichelberger H, et al. Association of body composition with survival and locoregional control of radiotherapy-treated head and neck squamous cell carcinoma. JAMA Oncol 2016;2:782–9.

[13] Fattouh M, Chang GY, Ow TJ, Shifteh K, Rosenblatt G, Patel VM, et al. Association between pretreatment obesity, sarcopenia, and survival in patients with head and neck cancer. Head Neck 2019;41:707–14.

[14] Chamchod S, Fuller CD, Mohamed ASR, Grossberg A, Messer JA, Heukelom J, et al. Quantitative body mass characterization before and after head and neck cancer radiotherapy: a challenge of height-weight formulae using computed tomography measurement. Oral Oncol 2016;61:62–9.

[15] Olson B, Edwards J, Degnin C, Santucci N, Buncke M, Hu J, et al. Establishment and validation of pre-therapy cervical vertebrae muscle quantification as a prognostic marker of sarcopenia in head and neck patients receiving definitive cancer surgery. MedRxiv 2021.

[16] Tajbakhsh N, Jeyaseelan L, Li Q, Chiang JN, Wu Z, Ding X. Embracing imperfect datasets: A review of deep learning solutions for medical image segmentation. Med Image Anal 2020;63:101693.

[17] Zhou T, Ruan S, Canu S. A review: Deep learning for medical image segmentation using multi-modality fusion. Array 2019;3:100004.

[18] Bakator M, Radosav D. Deep learning and medical diagnosis: A review of literature. Multimodal Technol Interact 2018;2:47.

[19] Naser MA, Deen MJ. Brain tumor segmentation and grading of lower-grade glioma using deep learning in MRI images. Comput Biol Med 2020;121:103758. https://doi.org/10.1016/j.compbiomed.2020.103758.

[20] Amarasinghe KC, Lopes J, Beraldo J, Kiss N, Bucknell N, Everitt S, et al. A deep learning model to automate skeletal muscle area measurement on computed tomography images. Front Oncol 2021;11.

[21] Kanavati F, Islam S, Arain Z, Aboagye EO, Rockall A. Fully-automated deep learning slice-based muscle estimation from CT images for sarcopenia assessment. ArXiv Prepr 200606432 2020.

[22] Pickhardt PJ, Perez AA, Garrett JW, Graffy PM, Zea R, Summers RM. Fully Automated Deep Learning Tool for Sarcopenia Assessment on CT: L1 Versus L3 Vertebral Level Muscle Measurements for Opportunistic Prediction of Adverse Clinical Outcomes. Am J Roentgenol 2021:1–8.

[23] Burns JE, Yao J, Chalhoub D, Chen JJ, Summers RM. A machine learning algorithm to estimate sarcopenia on abdominal CT. Acad Radiol 2020;27:311–20.

[24] Graffy PM, Liu J, Pickhardt PJ, Burns JE, Yao J, Summers RM. Deep learning-based muscle segmentation and quantification at abdominal CT: application to a longitudinal adult screening cohort for sarcopenia assessment. Br J Radiol 2019;92:20190327.

[25] Paris MT, Tandon P, Heyland DK, Furberg H, Premji T, Low G, et al. Automated body composition analysis of clinically acquired computed tomography scans using neural networks. Clin Nutr 2020;39:3049–55.

[26] Elhalawani H, Mohamed ASR, White AL, Zafereo J, Wong AJ, Berends JE, et al. Matched computed tomography segmentation and demographic data for oropharyngeal cancer radiomics challenges. Sci Data 2017;4:170077.

[27] Grossberg A, Elhalawani H, Mohamed A, Mulder S, Williams B, White A, et al. HNSCC [Dataset]. Cancer Imaging Arch 2020. https://doi.org/10.7937/k9/tcia.2020.a8sh-7363.

[28] Clark K, Vendt B, Smith K, Freymann J, Kirby J, Koppel P, et al. The Cancer Imaging Archive (TCIA): maintaining and operating a public information repository. J Digit Imaging 2013;26:1045–57.

[29] Prado CMM, Lieffers JR, McCargar LJ, Reiman T, Sawyer MB, Martin L, et al. Prevalence and clinical implications of sarcopenic obesity in patients with solid tumours of the respiratory and gastrointestinal tracts: a population-based study. Lancet Oncol 2008;9:629–35.

[30] Anderson BM, Wahid KA, Brock KK. Simple Python Module for Conversions between DICOM Images and Radiation Therapy Structures, Masks, and Prediction Arrays. Pract Radiat Oncol 2021.

[31] Naser MA, Wahid KA, van Dijk L V, He R, Abdelaal MA, Dede C, et al. Head and Neck Cancer Primary Tumor Auto Segmentation using Model Ensembling of Deep Learning in PET-CT Images. MedRxiv 2021.

[32] The MONAI Consortium. Project MONAI. Zendo 2020. http://doi.org/10.5281/zenodo.4323059.

[33] Wahid KA, Ahmed S, He R, van Dijk L V, Teuwen J, McDonald BA, et al. Evaluation of deep learning-based multiparametric MRI oropharyngeal primary tumor auto-segmentation and investigation of input channel effects: Results from a prospective imaging registry. Clin Transl Radiat Oncol 2022;32:6–14. https://doi.org/10.1016/j.ctro.2021.10.003.

[34] Taha AA, Hanbury A. Metrics for evaluating 3D medical image segmentation: analysis, selection, and tool. BMC Med Imaging 2015;15:1–28.

[35] Chargi N, Bril SI, Emmelot-Vonk MH, De Bree R. Sarcopenia is a prognostic factor for overall survival in elderly patients with head-and-neck cancer. Eur Arch Oto-Rhino-Laryngology 2019;276:1475–86.

[36] Meerkerk CDA, Chargi N, de Jong PA, van den Bos F, de Bree R. Sarcopenia measured with handgrip strength and skeletal muscle mass to assess frailty in older patients with head and neck cancer. J Geriatr Oncol 2021;12:434–40.

[37] Zwart AT, Becker J-N, Lamers MJ, Dierckx RAJO, de Bock GH, Halmos GB, et al. Skeletal muscle mass and sarcopenia can be determined with 1.5-T and 3-T neck MRI scans, in the event that no neck CT scan is performed. Eur Radiol 2021;31:4053–62.

