## Appendix for "Deep Learning Auto-Segmentation of Cervical Neck Skeletal Muscle for Sarcopenia Analysis Using Pre-Therapy CT in Patients with Head and Neck Cancer"

**Appendix A: Supplementary Methods**

***Segmentation model***

As shown in **Figure A1**, the network consists of four convolution blocks in the encoding and decoding branches and a bottleneck convolution block between the two branches. All convolution layers use a kernel size of 3 except one convolution layer in the bottleneck, which uses a kernel size of 1. The number of output channels for each convolution layer is shown above each layer in **Figure A1**. Each convolution block in the encoding branch comprises of a two-strided convolution layer and a residual connection containing a two-strided convolution layer and a one-strided convolution layer. In the bottleneck, the residual connection contains two one-strided convolution layers. In the decoding branch, each block contains a two-strided transpose convolution layer, a one-strided convolution layer, and a residual connection. Batch normalization and Parametric ReLU activation functions were used throughout the architecture. The CT image acted as a one-channel input to the model. The skeletal muscle (SM) segmentation mask was provided as a two-channel output in a one-hot encoding, with the first channel the probability map of the background and the second of the region of interest. The architecture shown in **Figure A1** is for a 3D ResUNet model. The number of channels in the convolution layers are 16, 32, 64, 128, and 256. The 3D ResUNet model was trained to segment the C3 section and used the 3D CT images and ground truth segmentation of the C3 section for training. A similar architecture was used for the 2D ResUNet model with all 3D convolution layers replaced with 2D convolutions. This model was used to segment the SM using the 2D CT image at the C3 section as input.

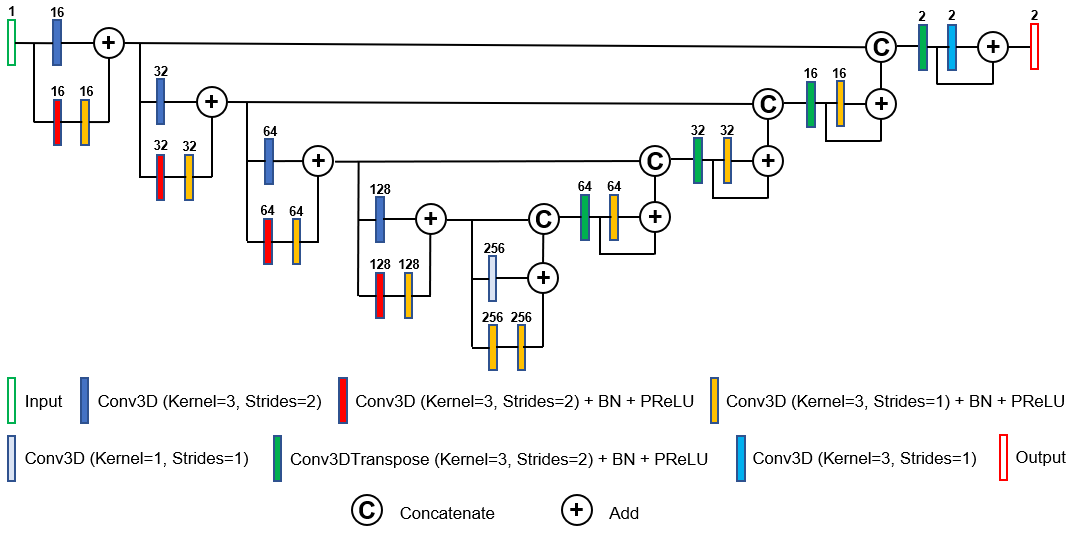

**Figure A1.** Schematic of the 3D ResUNet architecture used for the C3 section segmentation model. The number of channels is given above each block. The Batch normalization (BN) and the parametric ReLU (PReLU) layers are indicated. The 2D ResUNet model used for the SM segmentation has a similar architecture as shown in the figure with all Conv3D and Conv3DTranspose layers replaced with Conv2D and Conv2DTranspose layers, respectively.

***Model implementation***

For the 3D ResUNet model, the processed CT and C3 section masks were randomly cropped to four random fixed-sized regions (patches) of size (96, 96, 96) per patch per patient. The random spatial cropping considered the C3 section (i.e., the foreground) and non-C3 section (i.e., the background) in sampling, with a 50% probability for both the C3 and non-C3 sections. For a batch of training examples, we used 2 patients’ images and, therefore, a total of 8 patches of images. The shape of the input tensor provided to the network for an effective batch size of 8, a one-channel input (CT), and patch size of (96, 96, 96) was then, in channels-first format, (8, 1, 96, 96, 96). The C3 section mask was used as the ground truth target to train the segmentation model and its shape after the cropping process was (8, 1, 96, 96, 96). We implemented the same approach to train the 2D ResUNet model for SM segmentation. The shape of the input tensor provided to the network for a batch of 2 patients, patches per image of 4, a one-channel input (CT), and a patch size of (96, 96) is (8, 1, 96, 96). The SM segmentation mask was used as the ground truth target to train the 2D ResUNet model. The shape of the target tensor provided was (8, 1, 96, 96).
